# Plasma p-tau217 and incident mild cognitive impairment and dementia in older women: 25-year prospective study in The Women’s Health Initiative Memory Study

**DOI:** 10.1101/2025.10.30.25339146

**Authors:** Aladdin H. Shadyab, Bowei Zhang, Andrea Z. LaCroix, Michelle M. Mielke, Susan M. Resnick, Steve Nguyen, Luigi Ferrucci, Towia A. Libermann, Long Ngo, Ramon Casanova, Alexander P. Reiner, Danni Li, Caroline M. Nievergelt, Adam X. Maihofer, JoAnn E. Manson, Linda K. McEvoy

## Abstract

No study has evaluated whether associations of plasma phosphorylated tau 217 (p-tau217) with mild cognitive impairment (MCI) or dementia vary by race or hormone therapy (HT) use. We examined 2,766 cognitively unimpaired women ≥65 years randomized to HT vs placebo with 25-year follow-up. P-tau217 was associated with incident MCI/dementia (hazard ratio [HR], 2.43; 95% CI, 2.18-2.71) and each individual outcome (MCI: HR, 1.94; 95% CI, 1.72-2.20; dementia: HR, 3.17; 95% CI, 2.79-3.61). Associations between p-tau217 and dementia were stronger for women randomized to estrogen plus progestin vs placebo (HR, 4.18; 95% CI, 3.41-5.13 vs HR, 3.07; 95% CI, 2.41-3.91, respectively; *P* interaction=0.044) but did not vary for estrogen alone vs placebo. The combination of p-tau217 and age performed similarly in White and Black women (AUC=72.0% and 70.4%, respectively). Findings show the value of plasma p-tau217 for prediction of MCI and dementia up to 25 years in advance in older women.

---

Plasma biomarkers of Alzheimer’s disease (AD) offer a minimally invasive, accessible approach for detecting AD pathology.(1) Plasma phosphorylated tau 217 (p-tau217) has demonstrated higher accuracy in detecting AD pathology relative to other biomarkers,(2–5) with equivalent performance as cerebrospinal fluid p-tau217 for AD diagnosis.(6)

Some studies have found that plasma p-tau217 is associated with incident dementia.(7,8) However, several gaps in knowledge remain. Few studies have examined associations of plasma p-tau217 with risk of mild cognitive impairment (MCI) and dementia in community-based cohorts of cognitively healthy individuals with more than two decades of follow-up; prior studies had follow-up periods of 4-16 years.(7,8) Racial differences in the discriminative accuracy of plasma p-tau217 for MCI or dementia are not well understood, despite known disparities in dementia risk.(7–10) Notably, few studies have evaluated plasma p-tau217 in Black adults.(11–14) Although self-reported hormone therapy (HT) has been associated with increased tau accumulation and amyloid deposition as well as risk of dementia in observational studies,(15–19) no prior study has examined whether the association between plasma p-tau217 and cognitive outcomes differs by HT use.

The Women’s Health Initiative Memory Study (WHIMS) is the only large randomized clinical trial examining the effects of HT on cognitive outcomes among postmenopausal women.(20,21) In this study of 2,766 WHIMS participants, we examined associations of baseline plasma p-tau217 with incident MCI/dementia during up to 25 years of follow-up and evaluated whether associations varied by age, race, APOE ε4 genotype, or randomization to HT.

## METHODS

### Study design and population

WHIMS included two randomized controlled trials designed to investigate effects of HT on cognitive outcomes among 7,479 community-dwelling postmenopausal women ages 65-79 years who were cognitively unimpaired at randomization. Details on the WHIMS design and protocols are published.(20,21) Women were recruited from 39 US clinical centers during 1995-1998 and randomized either to oral conjugated equine estrogens (CEE) combined with medroxyprogesterone acetate (estrogen plus progestin; 2.5 mg/day) vs placebo among those with an intact uterus or oral CEE alone (estrogen alone; 0.625 mg/day) vs placebo among those with prior hysterectomy. The trials were stopped in 2002 and 2004, respectively; annual in-person follow-up continued through 2007. In 2008, WHIMS transitioned to annual telephone-administered cognitive assessments in the WHIMS-ECHO study, which followed participants for cognitive outcomes through 2021.(22) We examined 2,766 WHIMS women who were selected for biomarker measurements at randomization in 1995-1998 (see Supplementary Methods and Figure S1). This study was approved by the Institutional Review Board of University of California San Diego (Protocol No. 810929). All participants provided written informed consent. The WHIMS trials were registered at clinicaltrials.gov (ID: NCT00685009)

### Plasma p-tau217 measurement

Fasting blood was drawn at randomization and processed, frozen at −70°C, and shipped to a repository in Rockville, MD (Fisher Bioservices). Samples were shipped to the Advanced Research and Diagnostic Laboratory at University of Minnesota on dry ice for plasma p-tau217 measurement using the ALZpath Simoa pTau-217 v2 assay. Samples were assayed in singlets with the inclusion of 192 duplicates; laboratory personnel were blinded to the inclusion of these duplicates and to cognitive impairment status. The average intraassay coefficient of variation (CV) for plasma p-tau217 derived from duplicates was 11.4% (Supplementary Methods).

### Ascertainment of MCI and dementia

Our primary outcome was the combined endpoint of MCI or dementia; we also examined each individual outcome. MCI and dementia were ascertained and adjudicated annually through 2021 (Supplementary Methods). MCI diagnosis was based on Petersen’s criteria, and dementia diagnosis on *Diagnostic and Statistical Manual of Mental Disorders, Fourth Edition (DSM-IV)* criteria.(23,24)

### Covariates

Baseline questionnaires assessed age, education, smoking status, treated diabetes, cardiovascular disease, and total energy expenditure from recreational physical activity. Participants self-reported race (American Indian/Alaskan Native, Asian, Native Hawaiian/Other Pacific Islander, Black, White, more than one race, or unknown/not reported) and ethnicity (not Hispanic/Latino, Hispanic/Latino, or unknown/not reported). Hypertension was defined as either self-report of physician-diagnosed hypertension, use of hypertensive medications, systolic blood pressure ≥130 mm Hg, or diastolic blood pressure ≥80 mm Hg, measured at the clinic visit. Height and weight were measured to calculate body mass index (BMI; kg/m^2^). *APOE* ε4 carrier status was determined in White women only with available genome-wide genotyping data based on two single nucleotide variants, rs429358 and rs7412. Other covariates included HT treatment arm, high-density lipoprotein cholesterol (HDL), low-density lipoprotein cholesterol (LDL), and estimated glomerular filtration rate (eGFR), calculated based on serum creatinine measurements using the 2021 CKD-EPI equation.(25)

### Statistical Analysis

We excluded two outliers for p-tau217, defined as values more than five standard deviations above the mean. P-tau217 was log_2_ transformed and standardized (z-score). Continuous covariates were compared across quartiles of p-tau217 using Kruskal-Wallis tests. Categorical variables were compared using Chi-square tests. For variables with low expected counts, Fisher’s exact test with Monte Carlo stimulation with 2,000 replicates was used.

We generated weighted cumulative incidence curves for the outcomes according to p-tau217 quartiles, adjusting for the covariates above and accounting for competing risk of death. Weights were generated to account for the sampling design, defined as the product of inverse propensity score weights (IPW) and sampling weights (Supplementary Methods). We used weighted Cox proportional hazards regression models to determine hazard ratios (HRs) and 95% CIs for associations of p-tau217 with incident MCI/dementia. The 95% CIs were calculated using a robust sandwich variance estimator. Follow-up time was calculated from the date of WHIMS randomization, when p-tau217 was measured, to the date of the cognitive assessment that triggered the first diagnosis of MCI or dementia or the date of the final cognitive assessment (whichever came first), similar to prior WHIMS studies.(20,21) Models were adjusted for the covariates noted above according to prior literature.(26) We did not adjust for *APOE* ε4 genotype, as this variable was available in White women only, and adjustment did not materially change the findings; it was instead examined in subgroup analyses. The proportional hazards assumption was assessed using Schoenfeld residual plots and log-log survival plots; no violations were observed. To account for missing covariate data, we applied multiple imputation by chained equations using the R *mice* package, specifying all study variables with 20 imputations and 20 iterations. We repeated the above analyses using MCI and dementia as individual outcomes of interest.

In sensitivity analyses, we repeated the analyses accounting for competing risk of death using Fine-Gray regression models. We also evaluated risk of cognitive outcomes associated with quartiles of p-tau217 relative to the first quartile to identify potential non-linear associations. We excluded those with eGFR ≤ 60 mL/min/1.73 m², as chronic kidney disease may increase plasma biomarker levels.

In subgroup analyses, we examined whether associations with each outcome differed by assignment to HT. We also examined whether HT effects on MCI/dementia differed by baseline levels of p-tau217, using HT as the exposure of interest as in the original WHIMS trials and p-tau217 as the effect modifier. These latter analyses controlled for baseline eGFR, inverse probability weights, and sampling weights as described above but not for other covariates, given the randomized trial design. We next evaluated subgroup differences by race (Black vs White), age (≤70 years vs. >70 years), and *APOE* ε4 genotype (carrier vs. non-carrier). Differences by race were focused on Black and White women, given smaller sample sizes in other races. To assess potential effect modification, we included interaction terms between these factors and p-tau217 in the models.

We assessed the discriminative accuracy of p-tau217 using time-dependent receiver operating characteristic (ROC) curves generated from Cox models at the median follow-up. We determined sensitivity and specificity for p-tau217 alone and combined with demographic characteristics (age, race, and ethnicity). We present areas under the curve (AUC) in the overall sample and stratified by race.

All analyses were conducted using R version 4.4.3 (https://www.r-project.org/) in RStudio 2024.12.1 (https://cran.rstudio.com/). P-values were two-sided and considered significant at *P*<0.05.

## RESULTS

This study included 2,766 women selected for plasma p-tau217 measurement (Supplementary Figure S1). Differences between those included vs not included in the analytic sample are described in Supplementary Table S1. The mean (SD) baseline age was 69.9 (SD 3.8) years, 0.6% were American Indian/Alaskan Native, 4.5% were Asian, 18.0% were Black, 0.3% were Native Hawaiian or Other Pacific Islander, 74.0% were White, 2.8% were more than one race, 1.8% were unknown/not reported race, and 7.1% were Hispanic/Latina (Table 1). Women with higher levels of p-tau217 at baseline were more likely to be older, White, have lower BMI, have never smoked, have lower eGFR, be *APOE* ε4 carriers, and have higher HDL cholesterol; there were no other differences (Table 1). Differences in baseline characteristics between Black and White women are shown in Supplementary Table S2. Baseline plasma p-tau217 levels were lower on average in Black vs White women (Supplementary Figure S2).

**Table 1.**
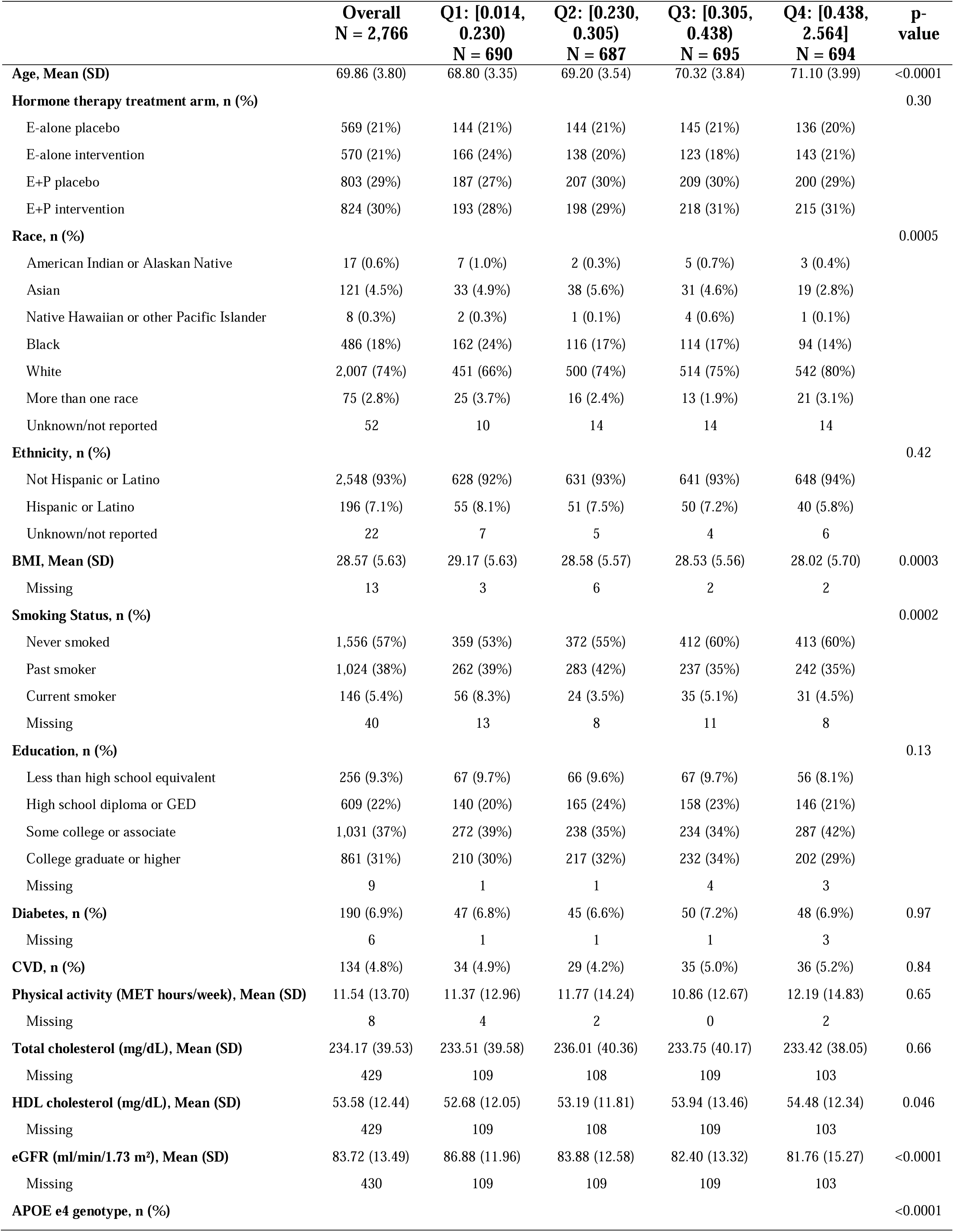
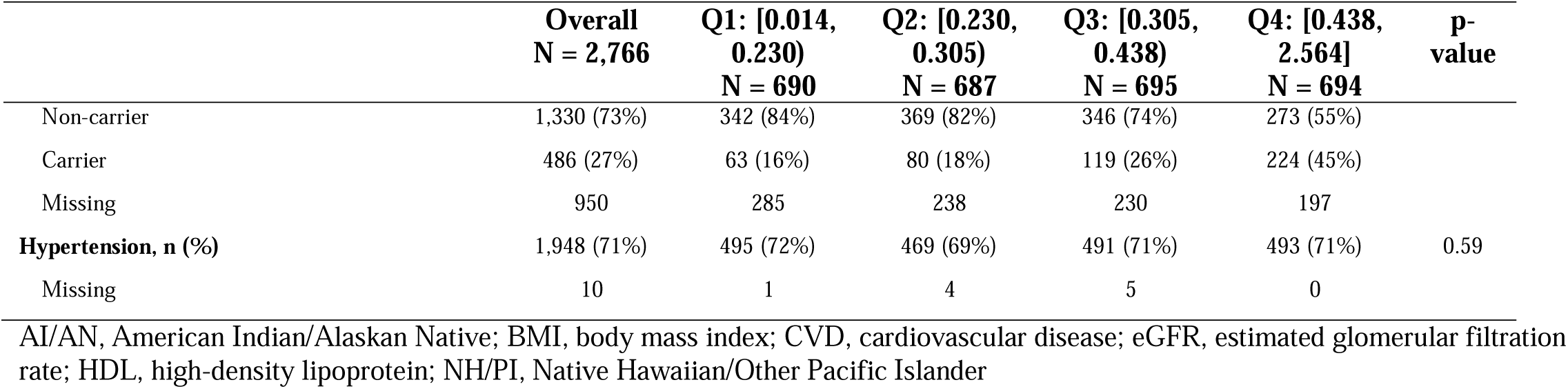
Baseline characteristics by baseline plasma p-tau217, Women’s Health Initiative Memory Study, 1995-1998.

During a mean (SD) follow-up of 13.39 (6.80) and median follow-up (IQR, range) of 14.05 (11.88; 0.88-25.15) years, 1,311 participants developed the combined endpoint of MCI/ dementia, with 849 cases of MCI and 752 cases of dementia. Cumulative incidence curves of MCI/dementia by quartiles of plasma p-tau217 are shown in Figure 1. Women in the highest quartile of p-tau217 showed a higher incidence of MCI/dementia shortly after baseline, a pattern observed in both White and Black women. For women in the third quartile, higher incidence of dementia became apparent after approximately 15 years, with little increase in dementia incidence for those in the first and second quartiles over the 25-year follow-up period.

**Figure 1.**
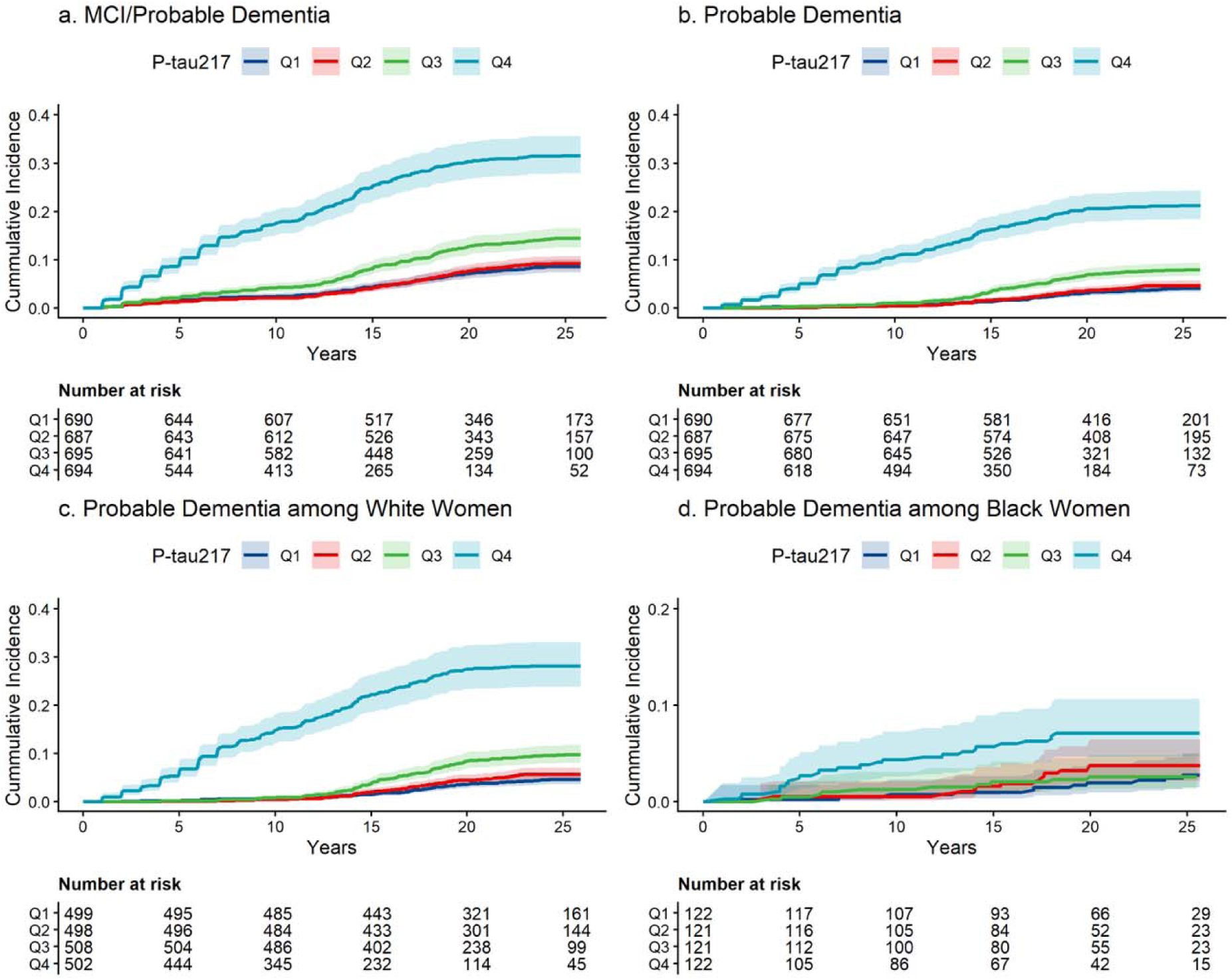
Weighted cumulative incidence curves of the combined endpoint of MCI/dementia and separately dementia across quartiles of baseline plasma p-tau217. Follow-up was capped at the 90^th^ percentile due to smaller risk sets from censoring leading to unstable survival estimates after 20 years of follow-up. Numbers at risk for unweighted analysis are shown below figure.

Associations of p-tau217 with the cognitive outcomes are shown in Figures 2-4. In the fully adjusted model, higher p-tau217 was associated with greater risk of MCI/dementia (HR, 2.43; 95% CI, 2.18-2.71; Figure 2). The HR was strongest for dementia (HR, 3.17; 95% CI, 2.79-3.61; Figure 3) and was lower in magnitude but remained significant when examining MCI (HR, 1.94; 95% CI, 1.72-2.20; Figure 2). We observed higher risk of the cognitive outcomes in the third and fourth quartiles of p-tau217, with the strongest HRs in the fourth quartile across all cognitive outcomes (e.g., HR, 7.37; 95% CI, 5.30-10.25 for Q4 vs Q1 for dementia; Supplementary Table S3).

**Figure 2.**
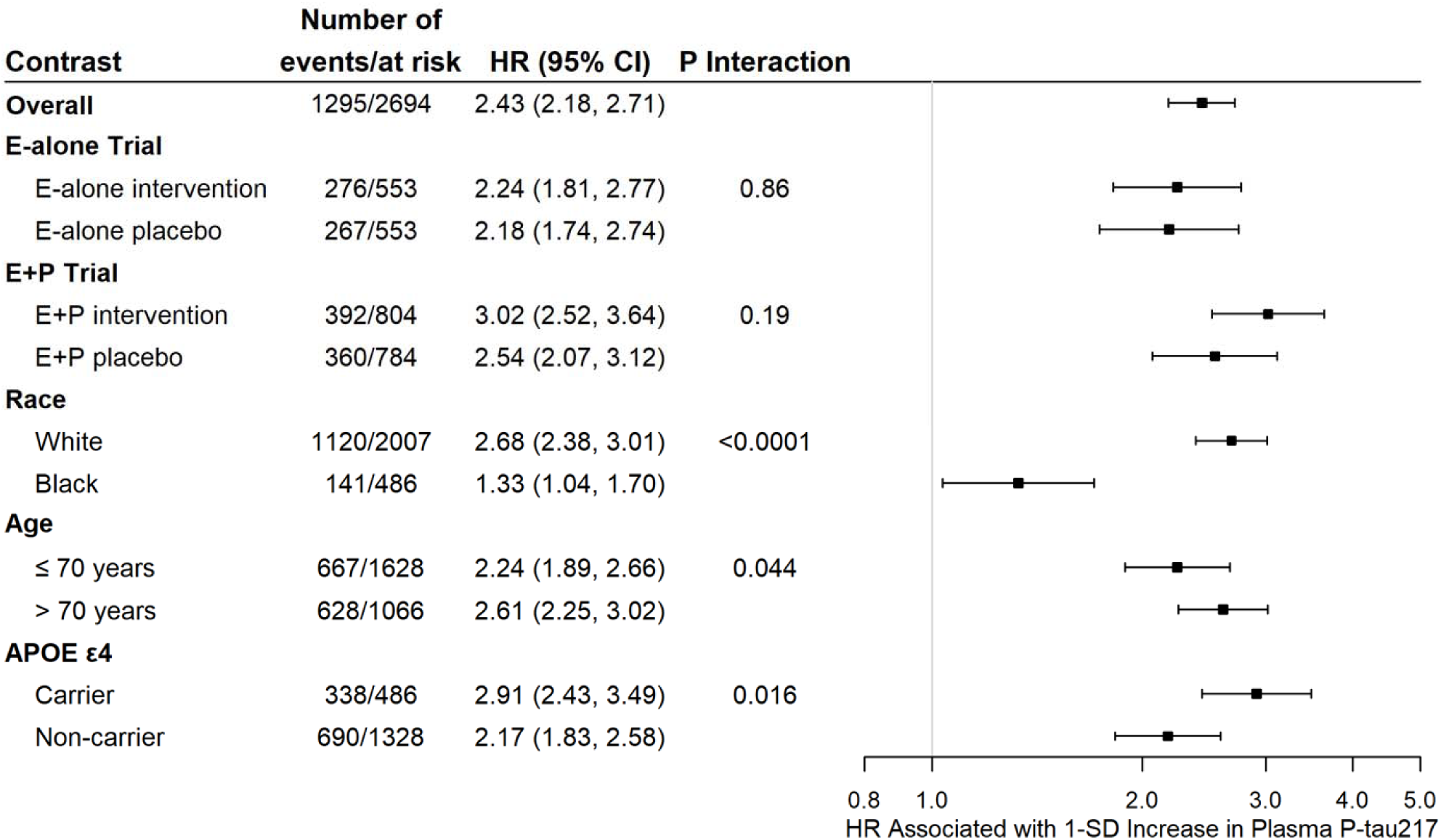
Association of baseline plasma p-tau217 with the combined endpoint of MCI/dementia overall and stratified by hormone therapy regimen, race, age, and *APOE* ε4. Hazard ratios (HRs) and 95% confidence intervals (CI) were derived from Cox proportional hazards regression models. All models were adjusted for hormone therapy trial arm, age, race, ethnicity, body mass index, smoking status, education, diabetes, cardiovascular disease, hypertension, physical activity, estimated glomerular filtration rate, total cholesterol, low-density lipoprotein cholesterol, high-density lipoprotein cholesterol, and inverse propensity weights and sample weights to account for sample selection. P-value for interaction modeled age as a continuous variable. The analytic sample included 2,694 women after removing those with unknown/not reported race. E-alone, estrogen alone; E+P, estrogen plus progestin.

**Figure 3.**
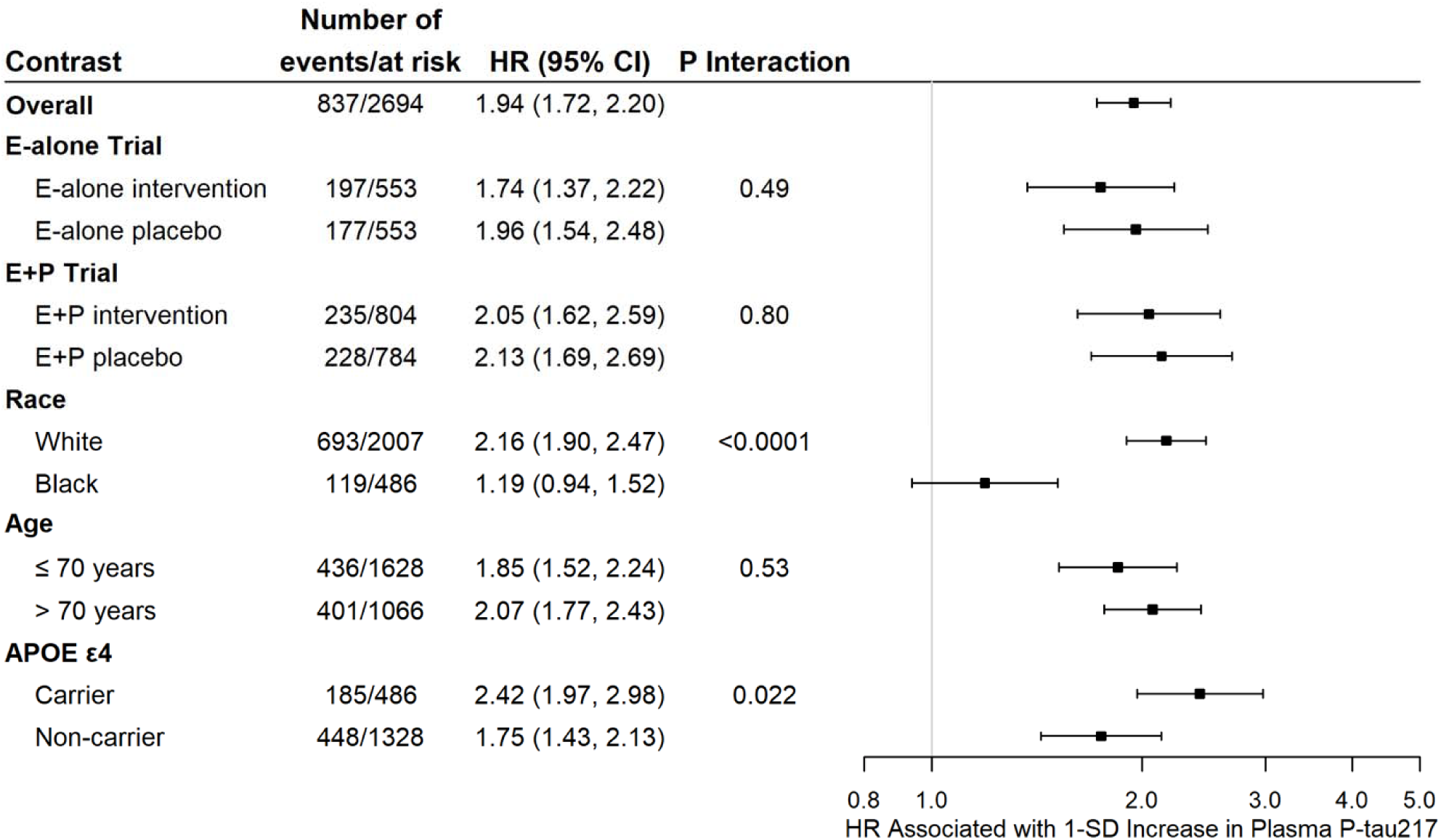
Association of baseline plasma p-tau217 with incident mild cognitive impairment overall and stratified by hormone therapy regimen, race, age, and *APOE* ε4. Hazard ratios (HRs) and 95% confidence intervals (CI) were derived from Cox proportional hazards regression models. All models were adjusted for hormone therapy trial arm, age, race, ethnicity, body mass index, smoking status, education, diabetes, cardiovascular disease, hypertension, physical activity, estimated glomerular filtration rate, total cholesterol, low-density lipoprotein cholesterol, high-density lipoprotein cholesterol, and inverse propensity weights and sample weights to account for sample selection. P-value for interaction modeled age as a continuous variable. The analytic sample included 2,694 women after removing those with unknown/not reported race. E-alone, estrogen alone; E+P, estrogen plus progestin.

**Figure 4.**
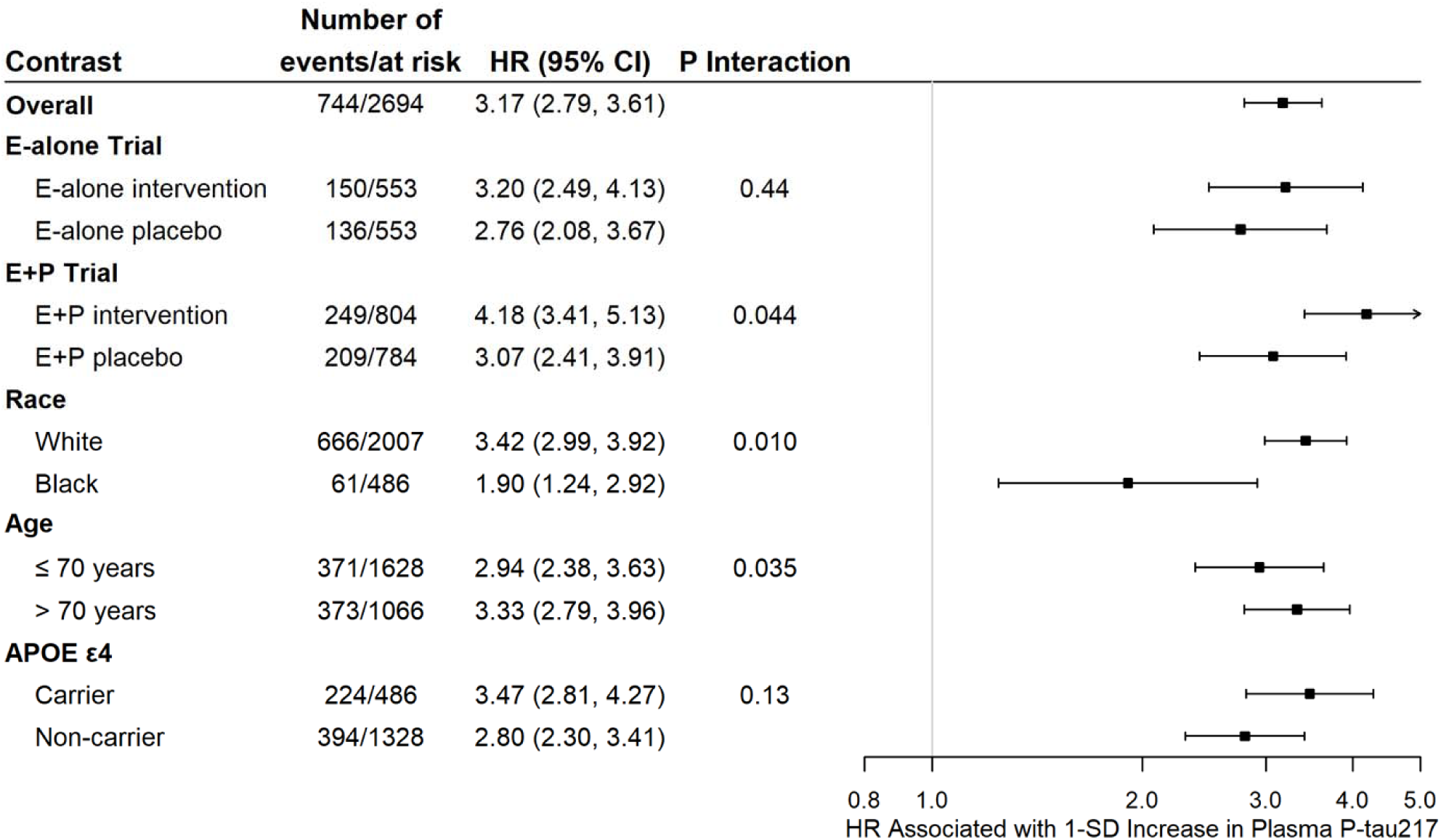
Association of baseline plasma p-tau217 with incident dementia overall and stratified by hormone therapy regimen, race, age, and *APOE* ε4. Hazard ratios (HRs) and 95% confidence intervals (CI) were derived from Cox proportional hazards regression models. All models were adjusted for hormone therapy trial arm, age, race, ethnicity, body mass index, smoking status, education, diabetes, cardiovascular disease, hypertension, physical activity, estimated glomerular filtration rate, total cholesterol, low-density lipoprotein cholesterol, high-density lipoprotein cholesterol, and inverse propensity weights and sample weights to account for sample selection. P-value for interaction modeled age as a continuous variable. The analytic sample included 2,694 women after removing those with unknown/not reported race. E-alone, estrogen alone; E+P, estrogen plus progestin.

We first examined whether the association between p-tau217 and cognitive outcomes differed by HT assignment at randomization, looking at each trial separately. The association of p-tau217 with cognitive outcomes did not significantly vary by estrogen alone vs placebo (Figures 2-4). Associations of p-tau217 with dementia were significantly greater in the estrogen plus progestin vs placebo group (HR, 4.18; 95% CI, 3.41-5.13 vs HR, 3.07; 95% CI, 2.41-3.91, respectively; *P* interaction=0.044; Figure 4); findings did not vary for the combined endpoint or MCI (Figures 2,3).

We next examined whether the effects of HT on MCI/dementia differed by baseline levels of p-tau217, wherein HT was a randomized exposure, as in the original WHIMS trials (Supplementary Figure S3). The risk of MCI/dementia (*P* interaction=0.90) and dementia separately (*P* interaction=0.68) did not vary for estrogen alone vs placebo across p-tau217 levels. There was a trend for increasing risk of dementia for estrogen plus progestin vs placebo with increasing p-tau217 (*P* for interaction=0.099) but not for the combined MCI/dementia endpoint (*P* interaction=0.28).

P-tau217 was more strongly associated with the cognitive outcomes among White vs Black women (Figures 2-4). P-tau217 was not associated with risk of MCI among Black women but was among White women (*P* interaction<0.0001; Figure 3). Associations of p-tau217 with the cognitive outcomes varied by *APOE* ε4, with stronger HRs in *APOE* ε4 carriers than non-carriers, an interaction that was significant for MCI/dementia (*P* interaction=0.016; Figure 2) and MCI (*P* interaction=0.022; Figure 3) but not dementia (*P* interaction=0.13; Figure 4). There were also subgroup differences by age, with stronger HRs for ages >70 vs ≤70 years for MCI/dementia (P interaction = 0.044; Figure 2) and dementia (*P* interaction=0.035; Figure 4) but not MCI (P interaction=0.53; Figure 3).

P-tau217 showed better discrimination than demographics for dementia prediction; the combination of p-tau217 and demographics outperformed either alone (AUC=72.7 %; Figure 5). In race-stratified analyses of dementia, p-tau217 alone had slightly higher performance in White vs Black women (Figure 5). The performance of the combination of p-tau217 and age was similar in White and Black women (AUC=72.0% vs AUC = 70.4%, respectively). AUCs for the combination of age and p-tau217 were higher in White vs Black women when examining MCI/dementia and MCI (e.g., 66.1% vs 57.3% for prediction of MCI, respectively; Supplementary Figures S4, S5).

**Figure 5.**
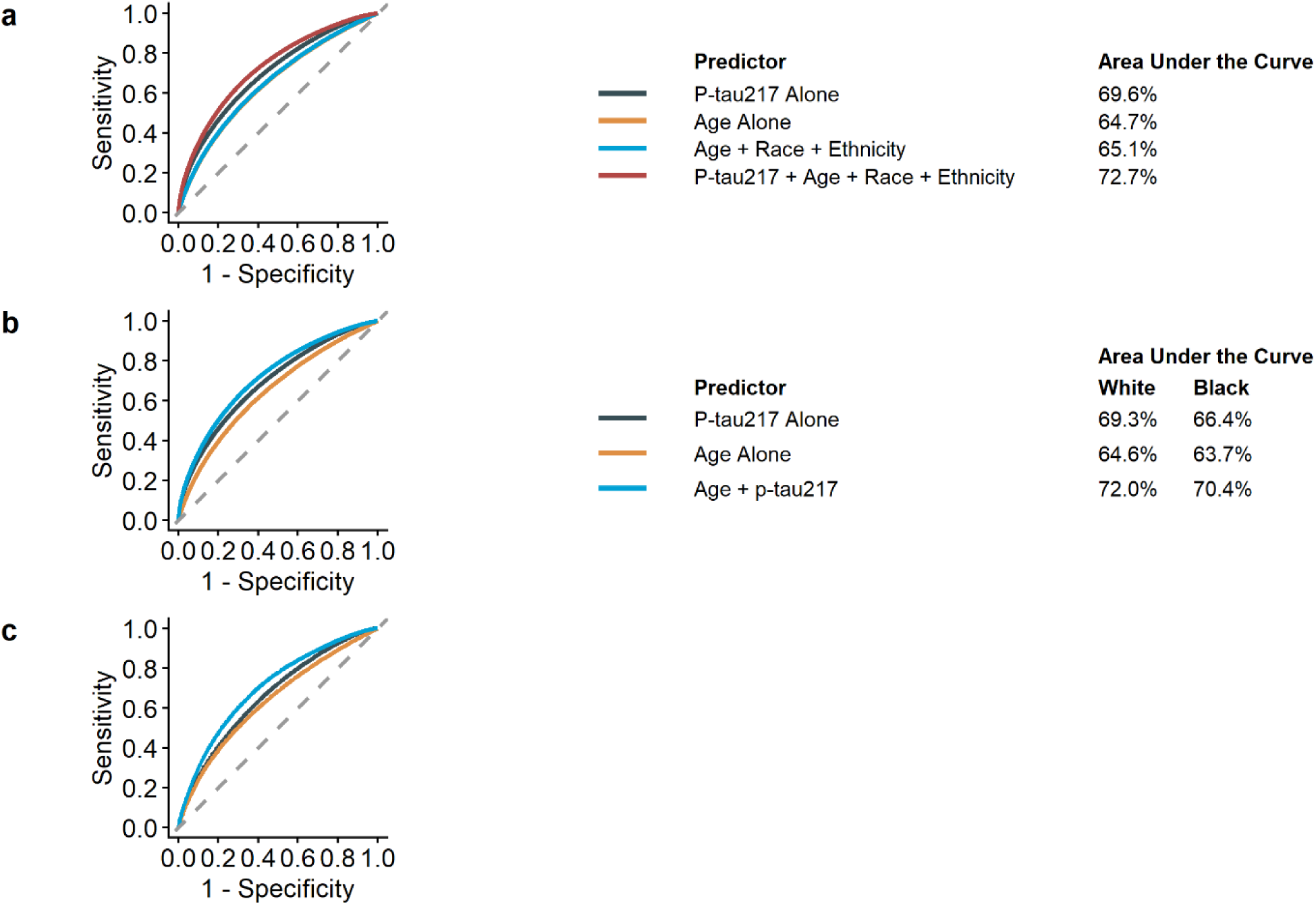
Discriminative accuracy of plasma p-tau217 for incident dementia examining (a) p-tau217 alone and in combination with age, race, and ethnicity in the overall sample; (b) p-tau217 among White women; and (c) p-tau217 among Black women. Receiver operating characteristic curves were generated from Cox proportional hazards regression models that estimated discriminatory accuracy at the median follow-up of 15.0 years in the full sample; 16.4 years in White women; and 8.0 years in Black women.

In Fine-gray regression models accounting for competing risk of death, associations of p-tau217 remained significant for all cognitive outcomes (Supplementary Figure S6). Findings were similar after excluding 524 women with eGFR ≤60 ml/min/1.73 m² (Supplementary Figure S7).

## DISCUSSION

In this prospective study among cognitively unimpaired postmenopausal women, we found that higher levels of plasma p-tau217 at baseline were associated with incident MCI and dementia during up to 25 years of follow-up. We show for the first time in the setting of a randomized controlled trial that higher plasma p-tau217 was associated with greater risk of dementia among women assigned to estrogen plus progestin compared with placebo. Plasma p-tau217 was more strongly associated with increased risk of MCI and dementia among White compared with Black women, although there was similar discriminative accuracy for dementia in both racial groups when combining p-tau217 with age (AUC, White: 72.0%; Black: 70.4%).

In the original WHIMS trials, estrogen alone compared with placebo increased risk of MCI/dementia by 38% (HR, 1.38; 95% CI, 1.01-1.89) but did not increase risk for each individual outcome,(21) whereas estrogen plus progestin compared with placebo doubled the risk of dementia (HR, 2.05; 95% CI, 1.21-3.48) but did not affect the risk of MCI (HR, 1.07; 95% CI, 0.74-1.55).(20) Here, we observed that higher levels of plasma p-tau217 at randomization were associated with greater risk of dementia for estrogen plus progestin vs placebo but not estrogen alone vs placebo.

We also observed a trend for higher dementia risk associated with estrogen plus progestin vs placebo by increasing baseline plasma p-tau217 levels but not for dementia risk associated with estrogen alone vs placebo. These findings suggest that risk of dementia associated with estrogen plus progestin may be higher among those with latent AD pathology and that risk of dementia associated with p-tau217 levels may differ by HT use.

There has been limited research on whether HT interacts with existing AD pathology to affect dementia risk. A study in 193 cognitively unimpaired women found that HT users with higher Aβ PET burden showed higher regional tau PET burden relative to non-users. Further, late initiation of HT (over 5 years after menopause) was associated with higher tau PET compared with early HT initiation (within five years of menopause).(15) However, this was a cross-sectional study of self-reported HT use; thus, causality cannot be determined.(15) Our findings in a randomized controlled trial provide evidence for the first time of a possible interaction between estrogen plus progestin and AD pathology related to increased dementia risk. It will be important to replicate these findings with other combination HT formulations.

We observed a three-fold higher risk of all-cause dementia with higher baseline p-tau217 levels at during a 25-year follow-up period. Among 435 cognitively unimpaired adults in the Swedish BioFINDER study, plasma p-tau217 was associated with two-fold higher risk of all-cause dementia during an average 4.8-year follow-up.(7) We also examined plasma p-tau217 as quartiles and found that the highest vs lowest quartile was associated with seven-fold increased risk of dementia. A study among 2,148 dementia-free adults (including those with MCI) from Sweden reported three-fold risk of all-cause dementia during the 16-year follow-up in those in the highest vs lowest quartile of baseline plasma p-tau217, with stronger findings in women vs men.(8) These findings support a potential dose-response relationship between p-tau217 and incident dementia, with markedly higher risk at the highest quartile.

We found that risk of MCI/dementia associated with p-tau217 levels was greater among women over 70 and *APOE* ε4 carriers. These findings are consistent with higher risk of MCI/dementia when p-tau217 levels are high in the presence of the most potent dementia risk factors, age and *APOE* ε4 carriage. A recent study among 2,148 dementia-free adults from Sweden observed higher risk of dementia for increasing plasma p-tau217 levels among APOE ε4 carriers vs non-carriers but reported that risk was higher among those who were <78 vs ≥78 years; in contrast to our study, the sample included both men and women as well as individuals with MCI.

Although the relationship between plasma p-tau217 and long-term risk of MCI has not been studied as extensively as risk of dementia, our results among White women are consistent with prior studies. A recent study among 1,474 cognitively unimpaired adults from nine cohorts reported that baseline plasma p-tau217 was associated with higher risk of incident MCI (HR, 1.57; 95% CI, 1.43-1.72) during a median 3.8-year follow-up.(27) The Mayo Clinic Study of Aging and BioFINDER-2 cohorts reported similar findings.(28) Our findings with a median follow-up of 14 years show that plasma p-tau217 may also be associated with longer-term risk of MCI.

Both White and Black women showed higher risk of dementia for higher levels of p-tau217, with stronger HRs in White vs Black women. We also found that the combination of age and p-tau217 showed better ability to discriminate women who developed dementia from those who did not, with similar AUCs between White and Black women (AUC=72.0% and 70.4%, respectively). However, there was no significant association of p-tau217 with incident MCI risk among Black women. None of the prior studies examined differences in p-tau217 associations with MCI risk by race. Both BioFINDER and the Mayo Clinic Study of Aging comprise primarily of White adults; the racial composition in the nine-cohort study described above was not provided. Although few studies have examined plasma p-tau217 associations with PET evidence of amyloid and tau pathology in Black adults, existing evidence suggests that plasma p-tau217 may be similarly sensitive to these brain pathologies in Black and White adults. A cross-sectional study among 233 Black adults (mean age, 64.5 years; 82% cognitively unimpaired) reported that plasma p-tau217 had high accuracy for detecting abnormal amyloid and tau PET (AUC=0.90 for amyloid PET and 0.89 for tau PET).(14) Another study in 289 older adults (28% Black; 62% cognitively normal) reported similar discriminative accuracy of plasma p-tau217 for amyloid positive status among White and Black adults.(12) The lack of association of plasma p-tau217 with risk of MCI among Black women in our study may thus indicate differences in MCI etiology between racial groups (e.g., vascular disease in Black women) or reflect misdiagnosis of MCI in Black women.(29,30)

Our study has several limitations. We examined only older women. We examined clinical diagnosis of dementia and did not have data on cerebrospinal fluid or neuroimaging biomarkers for diagnosis. Dementia subtyping was not performed in WHIMS. It is possible that the observed racial differences in biomarker prediction may arise from differences in comorbidities, underlying causes of dementia, and inequities in the accuracy of cognitive tests, some of which we were unable to address. The HT formulations in WHI may not be generalizable to currently favored HT regimens with lower doses, transdermal preparations, or micronized progesterone. Whether the findings can be generalized to HT initiated earlier in menopause is unknown. Finally, cautious interpretation of the nominally significant interactions in our study is warranted; while they have plausible explanations, they could also have occurred by chance.

Study strengths include the large, diverse sample of community-dwelling women with extensive follow-up. The WHIMS HT Trials had minimal exclusion criteria so that the study results would be maximally generalizable to postmenopausal women. Women were randomized to HT or placebo, providing the unique opportunity to assess whether associations of plasma p-tau217 with MCI/dementia differed by HT regimen, without the usual challenges invoked by self-selection and self-report of HT use.

Among a large sample of cognitively unimpaired, postmenopausal women, higher baseline plasma p-tau217 was associated with increased risk of MCI/dementia during up to 25 years of follow-up. Including plasma p-tau217 improved prediction of dementia relative to demographic characteristics alone, with similar performance in Black and White women. However, plasma ptau217 was not as predictive of MCI in Black women as in White women, suggesting a need to further investigate MCI as a diagnostic construct across racial groups. Plasma p-tau217 was associated with significantly greater risk of dementia among women assigned to estrogen plus progestin compared with placebo. This suggests that it may be important to take HT use into account when using plasma p-tau217 for risk stratification and dementia prediction. Our findings also suggest that other factors, such as age and *APOE* ε4, may be important to determine individualized risk of cognitive outcomes. While our results suggest that plasma p-tau217 is similarly useful for dementia prediction in White and Black women, future studies are needed to determine whether this is true for other racial and ethnic groups and men. Future studies are also needed to determine whether these findings apply to more recent HT regimens. Overall, these findings support the value of plasma p-tau217 as an easily measured biomarker for future MCI and dementia that may have a variety of uses in both research and clinical practice among diverse populations.

## Authors’ Contributions

A.H.S. and L.K.M. contributed to the study concept and design and obtained funding. A.H.S. and L.K.M. wrote the first draft of the manuscript. B.Z. conducted the statistical analyses. D.L. led the collection of plasma p-tau217 biomarker data. A.H.S., B.Z., and S.N. had full access to the data in the study. B.Z. and S.N. accessed and verified the data. All authors provided input to the interpretation of results, revised and approved the final manuscript, and had final responsibility for the decision to submit for publication.

## Data Sharing

De-identified data from the study and supporting documents can be made available after publication to researchers with investigator support, after approval of a proposal by the WHI Publications and Presentations Committee and with a signed data access agreement (see https://www.whi.org/doc/PP-policy.pdf). Please contact Aladdin Shadyab and Linda McEvoy.

## Acknowledgements

This study was funded by grants R01AG079149 to A.H.S. and L.K.M., R01AG075884 to A.P.R., and K99AG082863 to S.N. from the National Institute on Aging. The WHI Program is funded by the National Heart, Lung, and Blood Institute, National Institutes of Health, and U.S Department of Health and Human Services (R01 HL105065, 75N92021D00001, 75N92021D00002, 75N92021D00003, 75N92021D00004, and 75N92021D00005). This research was supported in part by the Intramural Research Program of the National Institutes of Health (NIH) to L.F. and S.M.R. The Women’s Health Initiative Memory Study was funded in part by Wyeth Pharmaceuticals. WHIMS-ECHO was funded by NIA HHSN271-2011-00004C. The contributions of the NIH author(s) are considered Works of the United States Government. The National Heart, Lung, and Blood Institute has representation on the Women’s Health Initiative Steering Committee, which governed the design and conduct of the study, the interpretation of the data, and preparation and approval of manuscripts. The findings and conclusions presented in this paper are those of the author(s) and do not necessarily reflect the views of the NIH or the U.S. Department of Health and Human Services.

We thank the WHI participants, staff, and investigators. The short list of WHI investigators can be found at: https://www-whi-org.s3.us-west-2.amazonaws.com/wp-content/uploads/WHI-Investigator-Short-List.pdf. The full list of WHI Investigators can be found at the following site: http://www.whi.org/researchers/Documents%2520%2520Write%2520a%2520Paper/WHI%2520Investigator%2520Long%2520List.pdf

