## Supplementary Material for "Plasma p-tau217 and incident mild cognitive impairment and dementia in older women: 25-year prospective study in The Women’s Health Initiative Memory Study"

**Supplementary Methods**

**Supplementary Figure S1. Study flow diagram**

**Supplementary Table S1. Baseline characteristics by inclusion in analytic sample, Women's Health Initiative Memory Study, 1995-1998**

**Supplementary Table S2. Baseline characteristics by race, Women's Health Initiative Memory Study, 1995-1998**

**Supplementary Figure S2. Violin plot of baseline plasma p-tau217 overall and by Black and White race**

**Supplementary Table S3. Associations of quartiles of baseline plasma p-tau217 levels with incident cognitive outcomes**

**Supplementary Figure S3. Hazard ratios examining the effects of estrogen alone vs placebo or estrogen plus progestin vs placebo on cognitive outcomes according to plasma p-tau217 levels at randomization**

**Supplementary Figure S4. Discriminative accuracy of plasma p-tau217 for the combined endpoint of mild cognitive impairment/dementia**

**Supplementary Figure S5. Discriminative accuracy of plasma p-tau217 for mild cognitive impairment**

**Supplementary Figure S6. Association of baseline plasma p-tau217 with incident cognitive outcomes,** ***accounting for competing risk of death using Fine-Gray regression models***

**Supplementary Figure S7. Association of baseline plasma p-tau217 at baseline with incident cognitive outcomes, after excluding 524 women with eGFR ≤60 ml/min/1.73 m²**

### SUPPLEMENTARY METHODS

*MCI and Dementia Adjudication*

Briefly, participants completed the Modified Mini-Mental State Examination, with those scoring below specific cut points (<80 for those with <8 years of education and <88 for those with ≥9 years of education) completing a modified Consortium to Establish a Registry for AD battery of neuropsychological tests and standardized tests in person. A physician with expertise in dementia diagnosis classified women as having no dementia, MCI, or probable dementia. MCI diagnosis was based on Petersen's criteria, and dementia diagnosis was based on *Diagnostic and Statistical Manual of Mental Disorders, Fourth Edition (DSM-IV)* criteria.[1,2] All data were sent to the WHIMS Clinical Coordinating Center for review and central adjudication of final diagnosis by a panel consisting of a neurologist, geriatric psychiatrist, and geropsychologist. WHIMS-ECHO used a common, validated protocol of telephone-based cognitive assessments and informant reviews, and a similar protocol to that of WHIMS for ascertainment and central adjudication of final diagnosis.[3]

*Calculation of weights*

IPW were calculated using the following variables in a logistic regression model in the full WHIMS dataset (N = 7479): participation in various WHI sub-studies, case-control status, age, region, race, ethnicity, smoking status, hormone therapy trial arm, CVD, diabetes, any non-melanoma cancer, depressive symptoms, hysterectomy, history of female hormone use, BMI, and hypertension. Case-control sampling weights were assigned based on the population prevalence of MCI/probable dementia and the ratio of cases to controls in the dataset.[4,5] This approach allows estimation of the conditional probability under the population distribution using case-control data.

*Sample Selection*

Among the 7,479 WHIMS participants, we first selected all 1,334 women with incident MCI or probable dementia through the end of follow-up on December 31, 2021. We next selected 1,502 controls who did not have MCI or probable dementia during follow-up, including 565 who were enrolled in WHIMS-ECHO. Controls in our sample included all women who had brain imaging data (n=519) and all women from underrepresented populations (including American Indian/Alaskan Native, Asian, Native Hawaiian/Other Pacific Islander, Black, more than one race, and Hispanic/Latina; n=707). We excluded participants with unknown time-to-event data (N=47), missing plasma p-tau217 data (N = 21), and two outliers more than five standard deviations above the mean for p-tau217, leading to a final analytic sample of 2,766 women. The Cox models were focused on women without missing race or ethnicity (N=2,694), as these variables were not imputed in the analysis.

*Plasma P-tau217 Measurement*

The following interassay laboratory coefficients of variation (CVs) were derived from an ARDL pooled sample and the two kit controls, which were run on every plate along with the samples: 11.4%, 11.2%, and 12.9% at mean concentrations of 0.75, 0.39, and 0.15 pg/mL for plasma p-tau217, respectively. The lower limit of detection (LOD) was 0·012 pg/mL, and the lower limit of quantification was 0·06 pg/mL. No values were less than the LOD in our analytic sample.

**Supplementary Figure S1. Study flow diagram**

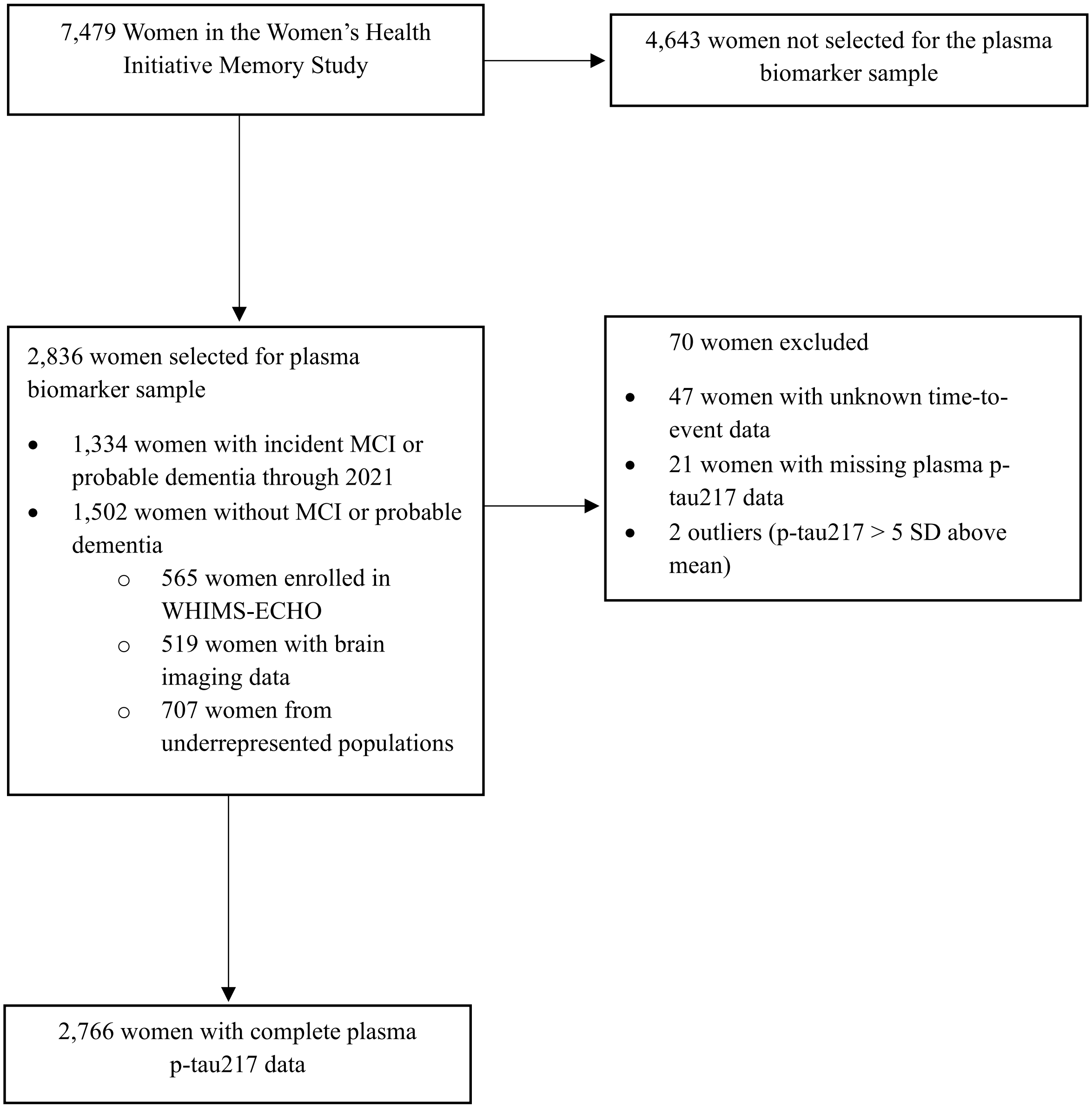

**Supplementary Table S1. Baseline characteristics by inclusion in analytic sample, Women's Health Initiative Memory Study, 1995-1998**

| | Analytic Sample (N = 2,766) | Not in Analytic Sample (N = 4,713) | Full WHIMS (N = 7,479) | p-value |
|---|---|---|---|---|
| **Age, Mean (SD)** | 69.86 (3·80) | 70.28 (3.86) | 70.12 (3.84) | <0.0001 |
| **Hormone therapy treatment arm, n (%)** | | | | 0.054 |
| Estrogen-alone placebo | 569 (21%) | 915 (19%) | 1,484 (20%) | |
| Estrogen-alone intervention | 570 (21%) | 899 (19%) | 1,469 (20%) | |
| Estrogen plus progestin placebo | 803 (29%) | 1,499 (32%) | 2,302 (31%) | |
| Estrogen plus progestin intervention | 824 (30%) | 1,400 (30%) | 2,224 (30%) | |
| **Race, n (%)** | | | | 0.0005 |
| American Indian or Alaskan Native | 17 (0.6%) | 0 (0%) | 17 (0.2%) | |
| Asian | 121 (4.5%) | 6 (0.1%) | 127 (1.7%) | |
| Native Hawaiian or other Pacific Islander | 8 (0.3%) | 0 (0%) | 8 (0.1%) | |
| Black | 486 (18%) | 37 (0.8%) | 523 (7.1%) | |
| White | 2,007 (74%) | 4,642 (99%) | 6,649 (90%) | |
| More than one race | 75 (2.8%) | 5 (0.1%) | 80 (1.1%) | |
| Unknown/not reported | 52 | 23 | 75 | |
| **Ethnicity, n (%)** | | | | <0.0001 |
| Not Hispanic or Latino | 2,548 (93%) | 4,685 (100%) | 7,233 (97%) | |
| Hispanic or Latino | 196 (7.1%) | 18 (0.4%) | 214 (2.9%) | |
| Unknown/not reported | 22 | 10 | 32 | |
| **BMI, Mean (SD)** | 28.57 (5.63) | 28.49 (5.74) | 28.52 (5.70) | 0.29 |
| Missing | 13 | 30 | 43 | |
| **Smoking Status, n (%)** | | | | <0.0001 |
| Never Smoked | 1,556 (57%) | 2,353 (51%) | 3,909 (53%) | |
| Past Smoker | 1,024 (38%) | 1,906 (41%) | 2,930 (40%) | |
| Current Smoker | 146 (5.4%) | 383 (8.3%) | 529 (7.2%) | |
| Missing | 40 | 71 | 111 | |
| **Education, n (%)** | | | | <0.0001 |
| Less than high school equivalent | 256 (9.3%) | 320 (6.8%) | 576 (7.7%) | |
| High school diploma or GED | 609 (22%) | 1,038 (22%) | 1,647 (22%) | |
| Vocational, training school, or some college or associate | 1,031 (37%) | 1,971 (42%) | 3,002 (40%) | |
| College graduate or higher | 861 (31%) | 1,371 (29%) | 2,232 (30%) | |
| Missing | 9 | 13 | 22 | |
| **Diabetes, n (%)** | 190 (6.9%) | 298 (6.3%) | 488 (6.5%) | 0.35 |
| Missing | 6 | 8 | 14 | |
| **Cardiovascular disease, n (%)** | 134 (4.8%) | 232 (4.9%) | 366 (4.9%) | 0.88 |
| **Physical activity (hours/week), Mean (SD)** | 11.54 (13.70) | 11.08 (13.04) | 11.25 (13.29) | 0.31 |
| Missing | 8 | 9 | 17 | |
| **Total cholesterol (mg/dL), Mean (SD)** | 234.17 (39.53) | 234.50 (40.30) | 234.38 (40.03) | 0.99 |
| Missing | 429 | 556 | 985 | |
| **HDL cholesterol (mg/dL), Mean (SD)** | 53.58 (12.44) | 53.29 (12.61) | 53.40 (12.55) | 0.28 |

| | Analytic Sample (N = 2,766) | Not in Analytic Sample (N = 4,713) | Full WHIMS (N = 7,479) | p-value |
|---|---|---|---|---|
| Missing | 429 | 556 | 985 | |
| **Hypertension, n (%)** | 1,948 (71%) | 3,321 (71%) | 5,269 (71%) | 0.96 |
| Missing | 10 | 11 | 21 | |
| **eGFR (ml/min/1.73 m²), Mean (SD)** | 83.72 (13.49) | 84.20 (13.49) | 84.03 (13.49) | 0.069 |
| Missing | 430 | 556 | 986 | |
| **APOE ε4 carrier status, n (%)** | | | | 0.073 |
| No ε4 alleles | 1,330 (73%) | 3,053 (75%) | 4,383 (75%) | |
| At least one ε4 allele | 486 (27%) | 994 (25%) | 1,480 (25%) | |
| Missing | 950 | 666 | 1,616 | |

Abbreviations: BMI, body mass index; CVD, cardiovascular disease; E-alone, estrogen-alone therapy; E+P, estrogen plus progestin therapy; eGFR, estimated glomerular filtration rate; HDL, high-density lipoprotein; mg/dL, milligrams per deciliter; ml/min/1.73 m², milliliters per minute per 1.73 square meters; SD, standard deviation

**Supplementary Table S2. Baseline characteristics by race, Women's Health Initiative Memory Study, 1995-1998**

| | Black<br>N = 486 | White<br>N = 2,007 | p-value |
|---|---|---|---|
| **Age, Mean (SD)** | 69.52 (3.74) | 69.92 (3.80) | 0.032 |
| **Hormone therapy treatment arm, n (%)** | | | <0.0001 |
| E-alone placebo | 139 (29%) | 374 (19%) | |
| E-alone intervention | 149 (31%) | 361 (18%) | |
| E+P placebo | 99 (20%) | 620 (31%) | |
| E+P intervention | 99 (20%) | 652 (32%) | |
| **Ethnicity, n (%)** | | | <0.0001 |
| Not Hispanic or Latino | 482 (99%) | 1,872 (93%) | |
| Hispanic or Latino | 4 (0.8%) | 135 (6.7%) | |
| **BMI, Mean (SD)** | 30.67 (6.10) | 28.25 (5.39) | <0.0001 |
| Missing | 4 | 8 | |
| **Smoking Status, n (%)** | | | <0.0001 |
| Never smoked | 240 (51%) | 1,153 (58%) | |
| Past smoker | 192 (41%) | 745 (38%) | |
| Current smoker | 41 (8.7%) | 86 (4.3%) | |
| Missing | 13 | 23 | |
| **Education, n (%)** | | | <0.0001 |
| Less than high school equivalent | 83 (17%) | 134 (6.7%) | |
| High school diploma or GED | 78 (16%) | 464 (23%) | |
| Some college or associate | 173 (36%) | 754 (38%) | |
| College graduate or higher | 149 (31%) | 649 (32%) | |
| Missing | 3 | 6 | |
| **Diabetes, n (%)** | 73 (15%) | 85 (4.2%) | <0.0001 |
| Missing | 2 | 4 | |
| **CVD, n (%)** | 39 (8.0%) | 76 (3.8%) | <0.0001 |
| **Physical activity (MET-hours/week), Mean (SD)** | 9.24 (13.30) | 12.03 (13.81) | <0.0001 |
| Missing | 2 | 5 | |
| **Total cholesterol (mg/dL), Mean (SD)** | 228.17 (45.26) | 235.43 (38.30) | 0.0004 |
| Missing | 126 | 103 | |
| **HDL cholesterol (mg/dL), Mean (SD)** | 55.72 (13.85) | 53.35 (12.15) | 0.0044 |
| Missing | 126 | 103 | |
| **eGFR (ml/min/1.73 m²), Mean (SD)** | 77.18 (16.33) | 84.85 (12.45) | <0.0001 |
| Missing | 126 | 104 | |
| **Hypertension, n (%)** | 408 (84%) | 1,339 (67%) | <0.0001 |
| Missing | 2 | 7 | |

Abbreviations: BMI, body mass index; CVD, cardiovascular disease; E-alone, estrogen-alone therapy; E+P, estrogen plus progestin therapy; eGFR, estimated glomerular filtration rate; HDL, high-density lipoprotein; mg/dL, milligrams per deciliter; ml/min/1.73 m², milliliters per minute per 1.73 square meters; SD, standard deviation

**Supplementary Figure S2. Violin plot of baseline plasma p-tau217 overall and by Black and White race. SD, standard deviation.**

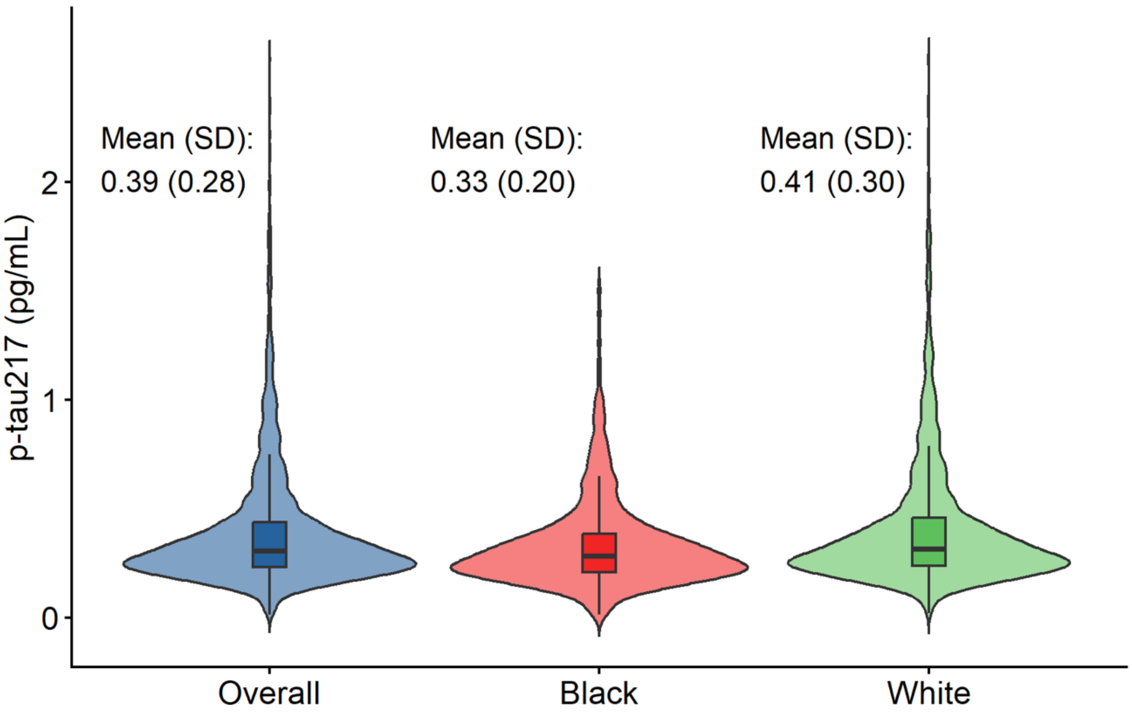

**Supplementary Table S3. Associations of quartiles of baseline plasma p-tau217 levels with incident cognitive outcomes**

| | Plasma P-tau217, pg/ml | HR (95% CI) |
|---|---|---|
| MCI/Dementia | 1st quartile: [0.014, 0.230) | 1 (reference) |
| | 2nd quartile: [0.230, 0.305) | 0.95 (0.73 - 1.25) |
| | 3rd quartile: [0.305, 0.438) | 1.44 (1.10 - 1.89) |
| | 4th quartile: [0.438, 2.564] | 4.69 (3.55 - 6.21) |
| MCI | 1st quartile: [0.014, 0.230) | 1 (reference) |
| | 2nd quartile: [0.230, 0.305) | 0.88 (0.66 - 1.17) |
| | 3rd quartile: [0.305, 0.438) | 1.36 (1.02 - 1.80) |
| | 4th quartile: [0.438, 2.564] | 3.30 (2.45 - 4.44) |
| Dementia | 1st quartile: [0.014, 0.230) | 1 (reference) |
| | 2nd quartile: [0.230, 0.305) | 1.06 (0.76 - 1.48) |
| | 3rd quartile: [0.305, 0.438) | 1.72 (1.24 - 2.37) |
| | 4th quartile: [0.438, 2.564] | 7.37 (5.30 - 10.25) |

CI, confidence interval; HR, hazard ratio; MCI, mild cognitive impairment

All models were adjusted for hormone therapy trial arm, age, race, ethnicity, body mass index, smoking status, education, diabetes, cardiovascular disease, hypertension, physical activity, estimated glomerular filtration rate, total cholesterol, low-density lipoprotein cholesterol, and high-density lipoprotein cholesterol. Models were also adjusted for inverse propensity weights and sampling weights to account for sample selection.

**Supplementary Figure S3. Hazard ratios examining the effects of estrogen alone vs placebo or estrogen plus progestin vs placebo on cognitive outcomes according to plasma p-tau217 levels at randomization.** Models adjusted for baseline eGFR as well as inverse probability and sampling weights. The sample sizes were 1,106 for the E-alone Trial and 1,588 for the E+P Trial. E-alone, estrogen alone; E+P, estrogen plus progestin.

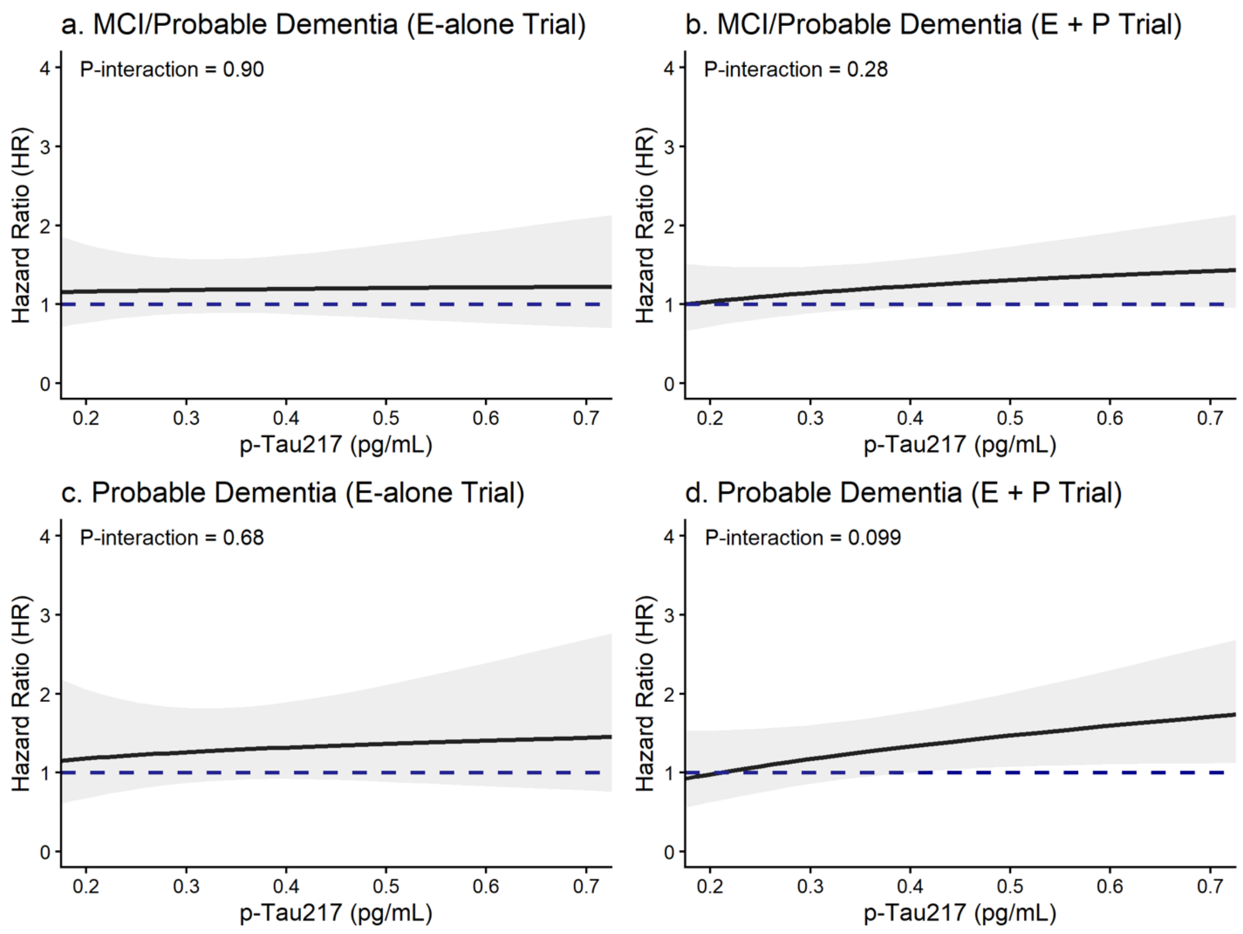

**Supplementary Figure S4. Discriminative accuracy of plasma p-tau217 for the combined endpoint of mild cognitive impairment/dementia examining (a) p-tau217 alone and in combination with age, race, and ethnicity in the overall sample; (b) p-tau217 among White women; and (c) p-tau217 among Black women.** Receiver operating characteristic curves were generated from Cox proportional hazards regression models that estimated discriminatory accuracy at the median follow-up of 14.1 years in the full sample, 15.4 years in White women, and 7.1 years in Black women. The sample included 2,694 women after removing those with unknown/not reported race.

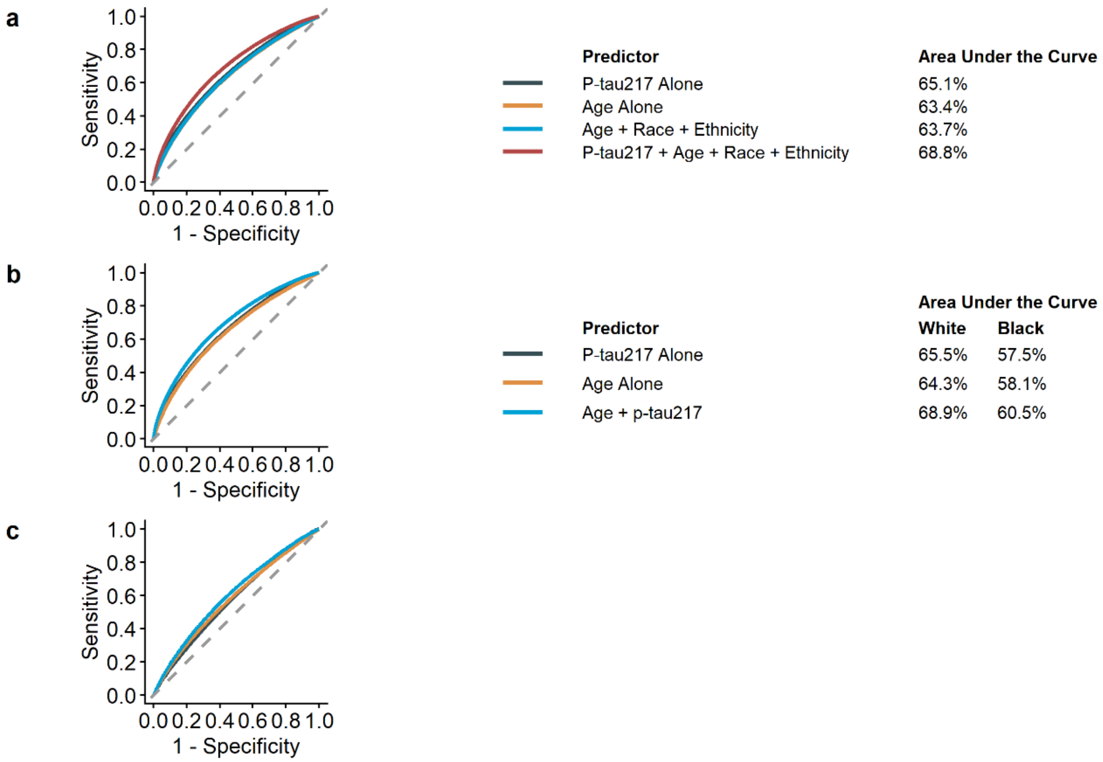

**Supplementary Figure S5. Discriminative accuracy of plasma p-tau217 for mild cognitive impairment examining (a) p-tau217 alone and in combination with age, race, and ethnicity in the overall sample; (b) p-tau217 among White women; and (c) p-tau217 among Black women.** Receiver operating characteristic curves were generated from Cox proportional hazards regression models that estimated discriminatory accuracy at the median follow-up of 14.1 years in the full sample, 15.7 years in White women, and 7.1 years in Black women. The sample included 2,694 women after removing those with unknown/not reported race.

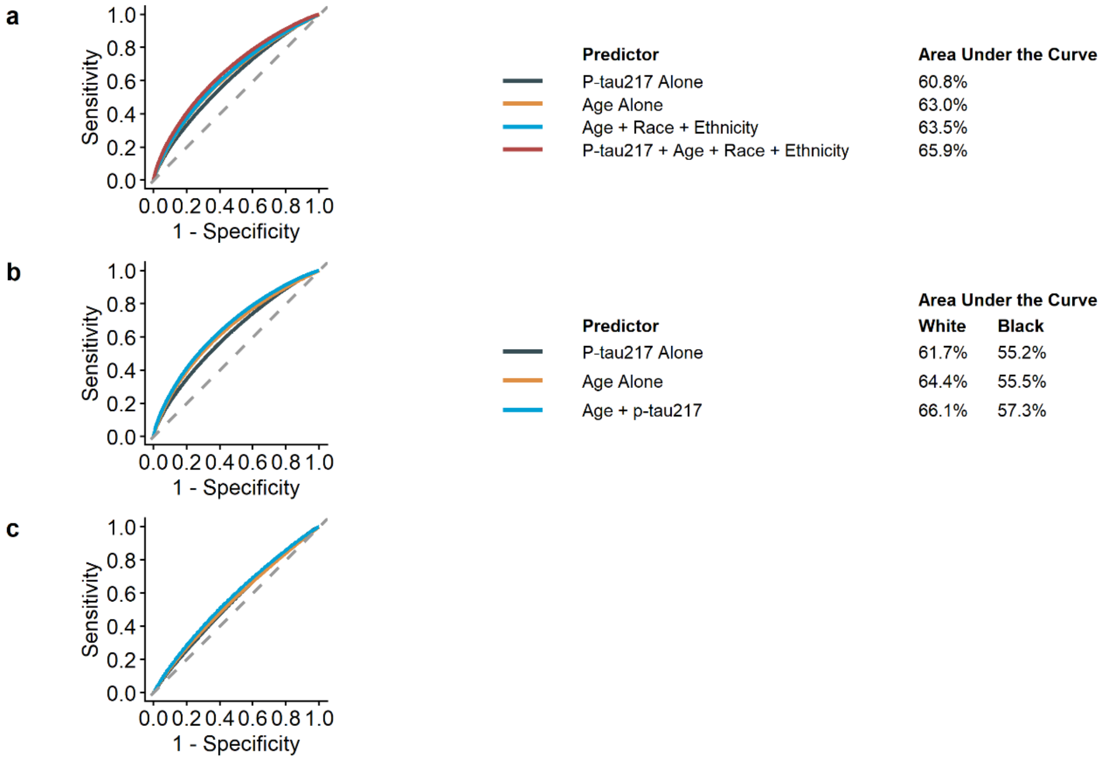

**Supplementary Figure S6. Association of baseline plasma p-tau217 with incident cognitive outcomes, *accounting for competing risk of death using Fine-Gray regression models*.** Subdistribution hazard ratios (SHRs) and 95% confidence intervals (CI) are shown. All models were adjusted for hormone therapy trial arm, age, race, ethnicity, body mass index, smoking status, education, diabetes, cardiovascular disease, hypertension, physical activity, estimated glomerular filtration rate, total cholesterol, low-density lipoprotein cholesterol, and high-density lipoprotein cholesterol. Models were also adjusted for inverse propensity weights and sampling weights to account for sample selection. The sample included 2,694 women after removing those with unknown/not reported race. MCI, mild cognitive impairment.

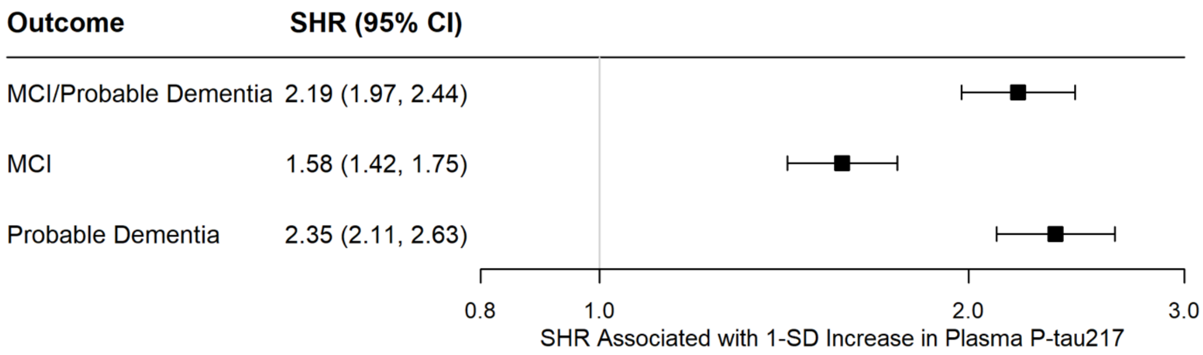

**Supplementary Figure S7. Association of baseline plasma p-tau217 at baseline with incident cognitive outcomes, *after excluding 524 women with eGFR ≤60 ml/min/1.73 $m^2$* (N=2,170).** Hazard ratios (HRs) and 95% confidence intervals (CI) were derived from Cox proportional hazards regression models. All models were adjusted for hormone therapy trial arm, age, race, ethnicity, body mass index, smoking status, education, diabetes, cardiovascular disease, hypertension, physical activity, estimated glomerular filtration rate, total cholesterol, low-density lipoprotein cholesterol, and high-density lipoprotein cholesterol. Models were also adjusted for inverse propensity weights and sampling weights to account for sample selection.

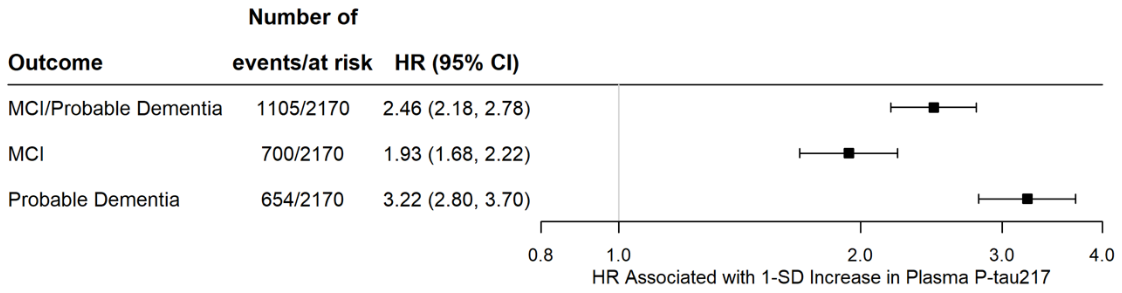
